# Estimating the malaria attributable fraction of fever in cohort studies through a before and after comparison of impact: Nagongera, Tororo, Uganda, 2011-2019

**DOI:** 10.1101/2022.12.29.22284050

**Authors:** John Rek, David Galick, Emily Hilton, John M. Henry, Austin Carter, Joaniter I. Nankabirwa, Emmanuel Arinaitwe, Maato Zedi, Paul Krezanoski, Isabel Rodriguez-Barraquer, Bryan Greenhouse, Robert C. Reiner, Simon I. Hay, Moses R. Kamya, Grant Dorsey, Jimmy Opigo, David L. Smith

## Abstract

**Background:** Malaria is an important cause of fever across much of sub-Saharan Africa and other places where *Plasmodium falciparum* infection is highly prevalent. Here, we estimate the fraction of fever that is attributable to malaria using data from two studies in Nagongera, Tororo, Uganda that followed cohorts of children and adults longitudinally from 2011-2019. The study included three years before and five years after indoor residual spraying (IRS) sharply reduced mosquito populations, malaria exposure, and the prevalence of malaria infection.

**Methods:** We estimate the malaria attributable fraction of fever (MAFF) by directly quantifying and comparing fever before and after IRS started. We compared subjective (*i*.*e*., self-reported) and objective fever during scheduled and unscheduled visits (*i*.*e*., to seek care) in young children (under 5 years old), older children (aged 5-10 years), and adults (over 18 years old).

**Results:** We estimated that there were 78-90 total days *per person, per year* (*pppy*) with subjective fever during the pre-IRS baseline in young children; 52-58 in older children; and 38-46 days in adults. After IRS, sub-clinical fever declined to 5-6 days *pppy* with fever in young children to around 3 in older children, and around 1 in adults: a 94% reduction in young children, 95% in older children, and 99% in adults. Reductions in total fever prevalence for care seeking (during unscheduled visits) declined by around 50% in young children, 65% in older children, and 80% in adults. In the before *vs*. after comparison, malaria accounted for 88% of objective fever during scheduled visits in young children, 75% in older children, and 91% in adults. Total fever declined by 80-85% in children and 90-93% in adults. During care seeking, malaria accounted for around 44% of objective fever in young children, but no meaningful differences were observed at other ages. These patterns were accompanied by changes in care seeking and total fever. Over the first few months of the study, care seeking rates increased in all groups, but then care seeking rates started a decline that continued until the study ended. By the end of the study, care seeking rates had declined by more than 75% overall compared with the start.

**Conclusions:** The fraction attributed to malaria differed by age and context. In this study population with good access to care, fever was rare at the end of the study in the absence of malaria. Based on the before vs. after comparison, malaria was directly or indirectly responsible for most subjective fever in the clinical setting, and it was also the dominant cause of objective fever. Surprisingly, a large fraction of subjective fever that occurred before IRS, during both scheduled and unscheduled visits, occurred in people who tested negative for malaria. The study draws attention to the importance of sub-clinical disease as a contributor to the burden of health in malaria endemic settings.

**Funding:** The PRISM studies (U19AI089674) were funded by the National Institutes of Allergies and Infectious Diseases (NIAID) as part of the International Centers of Excellence for Malaria Research (ICEMR).

## Background

Fever prevalence and malaria incidence are high across sub-Saharan Africa, partly because *Plasmodium falciparum* is highly endemic (D’Acremont et al., 2010; Dalrymple et al., 2017; Weiss et al., 2019; Maze et al., 2018). In areas where the overall incidence of disease is high, it is difficult to attribute disease symptoms and care seeking to any particular cause or to quantify interactions among multiple causes. An important question in sub-Saharan Africa and other areas where malaria is highly endemic is what fraction of fever is caused by infection with malaria parasites. In epidemiology, the fraction of disease attributable to a particular cause, called the etiological or population attributable fraction, is defined by the change in the incidence or prevalence of disease symptoms that would occur if that cause were removed (Levin, 1953; Rockhill et al., 1998). Ideally, the fraction of fever attributable to malaria would thus be measured by removing only malaria and comparing fever prevalence in populations with and without it, which has never been done (Koram and Molyneux, 2007; McGuinness et al., 1998; Rockhill et al., 1998). Recently, a pair of studies in Tororo, Uganda followed cohorts of individuals over a period of time when malaria reduced from a baseline of high exposure and high endemicity to very low levels through indoor residual spraying (IRS)(Nankabirwa et al., 2020). The studies included three years of pre-IRS baseline data, and five additional years of follow-up in a population with ongoing IRS. The IRS suppressed malaria transmission and reduced exposure to malaria, so we compared fever prevalence before and after an intervention to estimate the MAFF.

A core problem in malaria epidemiology has been how to define a malaria case (Koram and Molyneux, 2007). A concern has been that a substantial amount of fever is not attributable to malaria, even when the parasites are detected. For malaria in sub-Saharan Africa, *P. falciparum* infections can last for several months, during which time fever is intermittently present (Lindblade et al., 2013). The problem of defining the MAFF in malaria endemic regions is difficult because when parasite infections are highly prevalent fever arising from any cause could easily be misattributed to malaria. Most case definitions require both the presence of disease symptoms, usually including fever, and detection of malaria parasites. One method attributes fever statistically by correlating parasite densities estimated by microscopy with objective fever prevalence: the fraction of fevers attributed to malaria increases with parasite densities (Smith et al., 1994). Alternative methods based on a similar concept and underlying assumptions have also been proposed (Qin and Leung, 2005; Vounatsou et al., 1998; Wang and Small, 2012). The MAFF has also been estimated through associations between the prevalence of subjective fever and the prevalence of malaria (Dalrymple et al., 2017). Using these indirect methods, estimates of the MAFF for fever range widely depending on the study design, prevalence of malaria, and other contextual factors. Differences in studies estimating the MAFF have also differed in other ways, including attribution of subjective *vs*. objective fever, the population age, and attribution in cross-sectional studies *vs*. care-seeking patient populations (Afrane et al., 2014; Bisoffi et al., 2010; Bloland et al., 1999; Genton et al., 1995; Mabunda et al., 2009; Owolabi et al., 2016; Prybylski et al., 1999; Tchuinkam et al., 2015; Terrazas et al., 2015; Wang and Small, 2012). Ultimately, these indirect methods are based on assumptions that must be validated. There has long been a need for studies that measure the MAFF directly by observing fever after removing exposure to malaria.

A natural “experiment” occurred in Nagongera sub-county, Tororo District, Uganda during the PRISM studies, a pair of studies that followed cohorts of individuals from 2011 through 2019. No interventions were planned or funded by the study, but a decision was made by the national malaria control program to conduct mass indoor residual spraying in Tororo District, which began in December 2014 and continued through to the present day. The IRS coverage in the cohort households was estimated to be above 97% in the households enrolled in the study across all rounds. By all measures, malaria transmission began to decline sharply after the first round of IRS. Malaria fell sharply after that first round, and it continued to decline through the end of the study (Nankabirwa et al., 2020). The estimated annual entomological inoculation rate fell by roughly 553-fold, from approximately 238 to 0.43 infectious bites, per person per year. Malaria incidence fell by about 55-fold from 2.96 to 0.054 cases, per person, per year. Malaria parasite prevalence also declined; using positivity by any measure (*i*.*e*., positive either by light microscopy or by qPCR), prevalence fell 10-fold from 67.8% to 6.8%. The IRS thus appears to have suppressed transmission effectively through the end of the cohort studies. More recently, the effectiveness appears to have waned after the studies ended, and malaria transmission has been resurgent (Epstein et al., 2022). No other mosquito-transmitted pathogens have been identified in the clinic as a major cause of fever, and there were no other large-scale changes to health policies. Because of the PRISM study design, it was possible to estimate the MAFF directly by age, to compare the MAFF in and out of the clinic, and to compare some aspects of subjective and objective fever.

## Methods

### Cohort Studies

The PRISM 1 and PRISM 2 studies were conducted in Nagongera, Tororo District, Uganda from August 2011 through October 2019. Details of the PRISM studies and a summary of the data have been reported elsewhere (Kamya et al., 2015; Nankabirwa et al., 2020). In brief, in PRISM 1, all children older than 6 months and up to and including 10 years of age and a primary caregiver were enrolled starting in August 2011 and followed through September 2017 (Kamya et al., 2015). In PRISM 2, starting in October 2017, all household residents were enrolled and followed through October 2019.(Nankabirwa et al., 2020) The PRISM 1 cohorts included most of the households from PRISM 1. All members of the household were given a long-lasting insecticide-treated net, and each 28 days (or hereafter, once a month) a CDC light trap was placed in each enrolled household next to the bed of one of the children enrolled in the study and left overnight. Participants were seen at a dedicated study clinic at Nagongera for all clinical care needs. Participants were also requested to come to the clinic for scheduled visits. The study protocol for the frequency of the scheduled visits changed over the course of the two studies (Fig. 1). Children aged 10 and under had scheduled visits once every three months through November 2014, and then once every month starting in December 2014. Adults had scheduled visits once every three months through July 2016, and then once every month starting in August 2016. Scheduled monthly visits continued through PRISM 2.

**Fig. 1:**
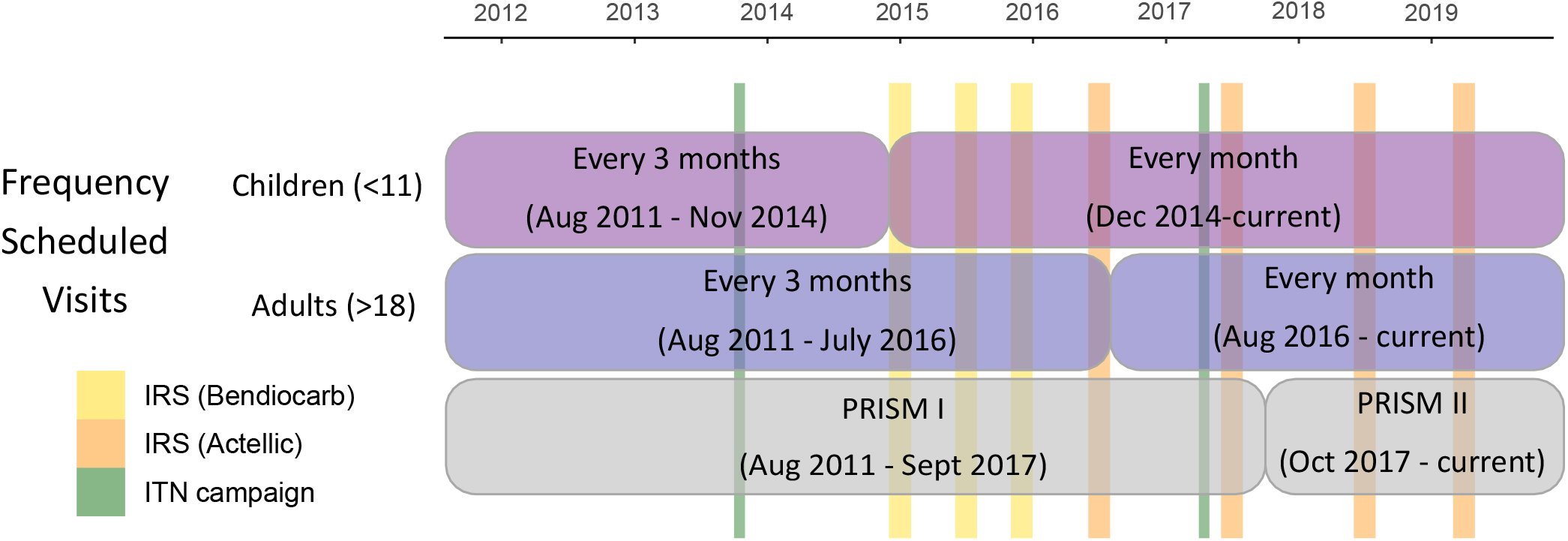
A timeline for PRISM 1 and PRISM 2 studies. Events illustrated include changes to the study protocol and mass distribution of vector control. ITN distributions occurred in October 2013 and March 2017 (green). IRS spray rounds started in Dec 2014, with three rounds of Bendiocarb (yellow; December 2014 - January 2015, June - July 2015 and November - December 2015), and four rounds of Actellic (Orange; June - July 2016, June - July 2017, June - July 2018 and March - April 2019). PRISM 1 enrolled only children aged 10 and under, and adults over the age of 18, and all participants had scheduled visits once per month until December 2014, when the frequency of scheduled visits for children increased to once per month (light purple). Adults continued having one scheduled visit every 3 months until August 2016, when they also increased in frequency to once per month (dark purple). The PRISM 2 study began in October 2017, allowing enrollment of all ages, and all participants had scheduled visits every month. Measurements of submicroscopic parasitemia at scheduled visits were made using LAMP in PRISM 1, and PCR in PRISM 2.

At all visits at the clinic, whether scheduled or not, participants had a detailed standard clinical evaluation, including assessment of history of fever in the last 24 hours, also called a subjective fever. During scheduled visits, temperature was also measured, and objective fever was defined as a documented temperature ≥38°C. Those patients with any fever during an unscheduled visit also had an urgent thick blood smear read for diagnosis of malaria. If diagnosed with malaria, they were prescribed antimalarial drugs according to the national guidelines and sent home. Regardless of fever status, a blood smear was taken during scheduled visits and read for malaria. Additionally, in scheduled visits submicroscopic parasitemia was measured by LAMP (PRISM 1) or qPCR (PRISM 2).

All individuals in the study were given a long-lasting insecticide treated net upon enrollment, which were replaced as needed throughout the study. During the study, government sponsored mass-distributions of LLINs occurred in October 2013 and March 2017. Government sponsored IRS spray programs occurred starting December 2014. Initially, three rounds of IRS with bendiocarb occurred approximately six months apart. These were followed by a gap of about six months before four additional rounds of IRS with Actellic^®^ occurred spaced about one year apart (Fig. 1).

Ethical approval for these studies were obtained from Makerere University, School of Medicine - Research and Ethical Committee, University of California, San Francisco Committee on Human Research Protection, London School of Hygiene and Tropical Medicine Ethics Committee and Uganda National Council for Science and Technology.

### Analysis Plan

In the analysis, scheduled and unscheduled visits were treated differently. We assumed that scheduled visits describe what is happening in the community, when there is no care seeking, so we call the proportion febrile during a scheduled visit the *community* or *sub-clinical fever* prevalence, meaning that it was not associated with an unscheduled visit to seek care. We call the proportion febrile during unscheduled visits the *clinical fever* prevalence, meaning that it was associated with a clinical visit to seek care. Community/sub-clinical fever and clinical fever are analyzed separately and discussed as distinct features of malaria epidemiology. A combined measure was developed to estimate the total number of fever days, per person, per year (*pppy*).

Subjective or objective fever during scheduled visits were summarized as a point estimate of prevalence, and since the day of the visit had been scheduled, we compared them to what would be found in a cross-sectional survey. Unscheduled visits in which subjects sought care in response to disease symptoms, were associated with overall disease incidence including malaria or any other cause of disease. Clinical fever prevalence was defined herein as the proportion of unscheduled visits where individuals had any fever. A clinical incident was defined by care seeking, or in other words, an unscheduled visit.

We used a simple model to add up total fever days, the number of days with fever, per person, per year. The total number of fever days, per person, per year in a population (denoted *F*) is a sum of two quantities. First, days with fever in the community, denoted *B*, was community fever prevalence times 365 days. Next, days with clinical fever was a product of three quantities: clinical incidence (the number of unscheduled visits, *pppy*, denoted *λ*); clinical fever prevalence (the proportion of incidents reporting with fever denoted *C*), and the average number of days with fever associated with each clinical incident with fever (*D*):

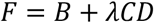

For example, 4 unscheduled visits in a year, of which 75% were febrile, and each causing 5 days of fever would be 15 days with a clinical fever. The number of days with fever per clinical incident was not known, so we conducted a sensitivity analysis using 1-10 days of fever per incident. We report the results, assuming each fever accompanying a clinical visit would last 3 days. If there were more days with fever associated with a febrile unscheduled visit, there would be a lower attributable fraction, and with fewer days per visit, a higher attributable fraction.

We report patterns in total fever days, community / sub-clinical fever prevalence, and clinical fever prevalence before and after the intervention. We also plotted these measures binned quarterly and aggregated by age: young children (under 5 years old), older children (aged 5-10 years), and adults (over 18 years old). Measures of mean fever prevalence and a binomial confidence interval were computed for each time-period. To estimate clinical incidence rates for each quarter, the numerator was the total number of unscheduled visits, and the denominator was the total number of unique individuals who were observed at least once during any reported period.

To compute the MAFF directly, as a before vs. after comparison, we defined two follow-up periods. The first follow-up period was from after the end of the third round of IRS (approximately one-year after the first round of IRS) through the end of the study, and the second follow-up period was for the last two years of the study. We computed total fever days under baseline conditions (denoted *F*_0_) to the total fever days during a follow-up period (denoted *F*_*i*_) using the formula:

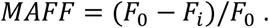

## Results

### Care Seeking Rates

To understand overall fever patterns, we first quantified the total number of scheduled and unscheduled visits, per person. The predominant patterns in the total number of scheduled and unscheduled visits were examine and summarized as a rate: the number of visits, per person, per quarter over the course of the study. Visits per quarter were rescaled to visits per year for figure labels.

Care seeking patterns were explained by several factors, including some that were related to the study design (Fig. 2). Enrollment was concentrated in the first month of PRISM 1 and the first three months of PRISM 2. Since the PRISM 1 protocol initially called for a scheduled visit once every three months, the enrollment process initiated a three-month cycle, where unscheduled patient visits peaked in the two months when a scheduled visit did not occur. This pattern persisted until the PRISM protocol changed to monthly scheduled visits. After the protocol changed to monthly visits for children, the frequency of scheduled visits increased sharply in children, but it fell short of targets until after the protocol changed for adults.

**Fig. 2:**
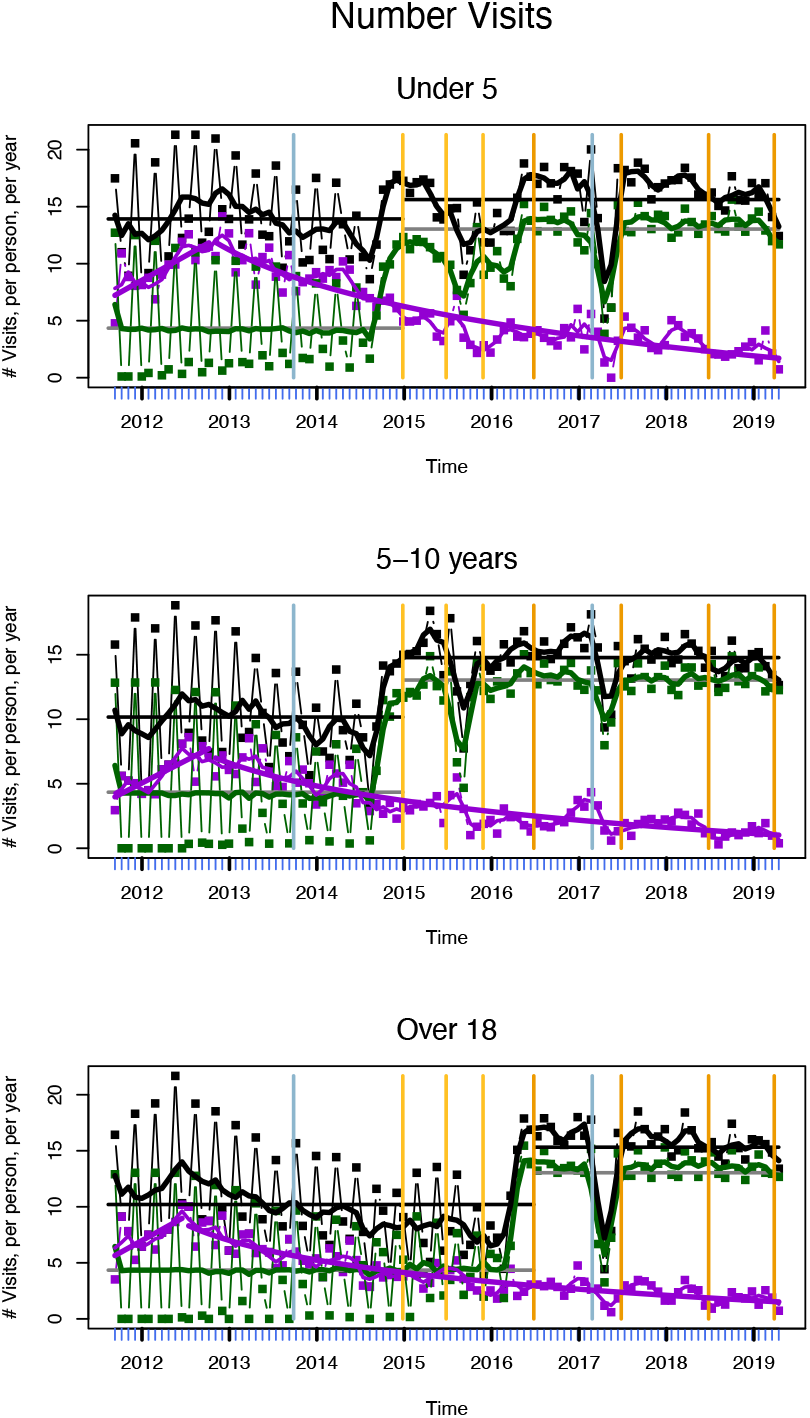
Care Seeking Rates and Patterns. The plots show care seeking rates, the number of scheduled (green), unscheduled (purple), and total visits (black), per person, per year, or *pppy*. The data are plotted here by month to highlight features of the study design (blue tick marks), but the rates are rescaled and reported per year. a) All ages; b) young children (<5); c) older children (5-10); and d) adults (>18). For each type of visit, we also show a smoothed trend line. Purple lines also show a fitted model: a linear trend line fitted to the first few months, and an exponential trend line fitted to the remainder. The break point was chosen by minimizing the difference. The average total numbers of visits *pppy* before and after the protocol change is plotted as solid black line segments. The protocol target for scheduled visits before and after the protocol change were plotted as grey line segments. The timing of vector control is shown with vertical lines (ITN in blue; IRS in yellow / orange), and the PRISM policy for scheduled visits occurs at the discontinuity in the grey line segment.

The frequency of unscheduled visits, meanwhile, increased to a peak sometime around the first year of the study. After peaking, the frequency of unscheduled visits declined through the end of the study. To identify the peak of care seeking, we defined several putative break points around the peak frequency, and then we fit a linear model to incidence vs. time before each break point, and a linear model to incidence vs. log(time) after each break point. A “best” break point was selected for analysis that minimized the difference in the values of the piecewise-defined model fits. The best break point was at 15 months, for young children, and at 11 months for older children and adults.

The care seeking rate rose from 5.6 visits up to around 8.4 visits, *per person, per year* (*pppy*) overall, before falling to 1.3 visits *pppy*. In young children, the rate rose from 7 visits *pppy* up to around 11.1 visits *pppy* around the 15th month of the study before falling to 1.6 visits *pppy*. In older children, there were 3.9 visits *pppy*, which rose to around 6.7 visits *pppy* around the 15th month of the study before falling to 1 visit *pppy*. In adults, there were 5.5 visits *pppy*, which rose to more than 8 visits *pppy* around the 11th month of the study before falling to 1.4 visits *pppy* (Fig. 2). Overall, trends / patterns in care seeking rates (*i*.*e*., unscheduled visits) did not show clear signs of changing after the mass distribution of ITNs, after IRS rounds, or after changes in the PRISM 1 study protocol. We considered these trends to be a long-term effect of improved access to care: healthcare utilization initially increases (increasing purple line in Fig. 2), followed by a decrease in utilization as health improves (decreasing purple line in Fig. 2).

The total number of visits *ppopy*, was affected by a study protocol change in which the frequency of scheduled visits shifted from once every three months to once every month. The lowest frequency of total visits occurred just before the study protocol changed. Not surprisingly, the overall frequency of visits increased after the protocol changed. The average number of visits, *pppy*, changed with the protocol from 12.9 to 14.7 visits *pppy* for young children (a 14% increase), from 9.3 to 13.8 visits *pppy* for older children (a 48% increase); and from 9.4 to 14.3 visits *pppy* for adults (a 52% increase).

## Fever Before vs. After IRS

### Fever Prevalence During Scheduled Visits

Community fever prevalence is defined herein as the fraction that reported a fever (subjective) or that had a measured fever (objective) during the scheduled visit, which included the enrollment visit and all subsequent regularly scheduled visits. Here, we report the analysis for any fever (Fig. 3) and then we focus on objective fever.

**Fig. 3:**
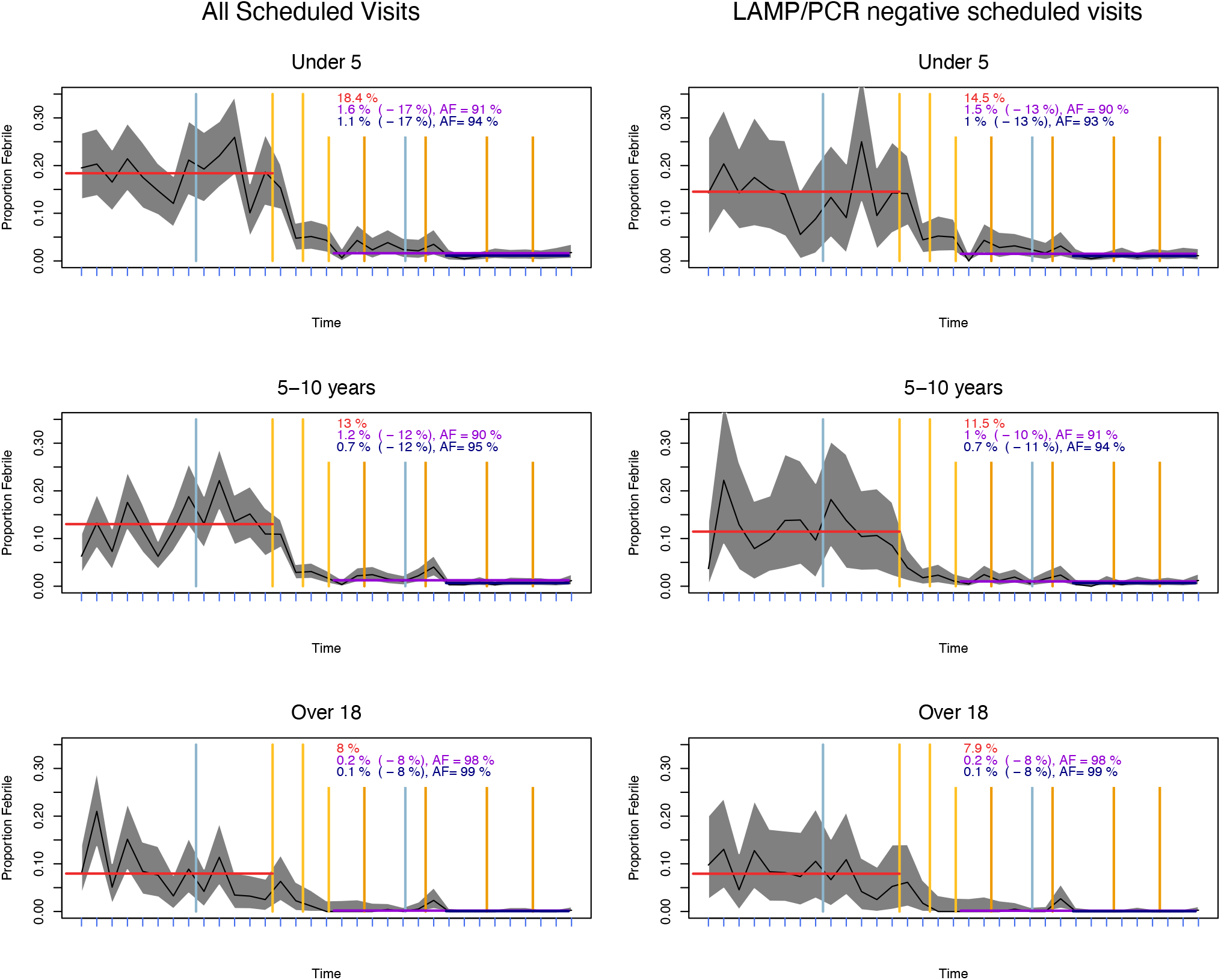
Community fever prevalence during scheduled visits. The data were binned and plotted by quarter (blue tick marks) for all scheduled visits (left), and for a subset of scheduled visits in which the subjects tested malaria-negative by light microscopy, LAMP (PRISM 1), and PCR (PRISM 2). The rows show young children (<5 years of age, top row), older children (5-10, middle row), and adults (>18, bottom row). Note that LAMP or PCR was conducted for 98% of scheduled visits. The shaded grey regions show the binomial confidence intervals. Vertical yellow/orange lines indicate dates when spraying occurred, and blue lines are the dates of mass ITN distribution. The red horizontal line and red text describe fever prevalence in the community at the pre-IRS baseline. The purple horizontal lines and text describe fever prevalence in the first follow-up period, the crude change in prevalence (in parenthesis), and the attributable fraction (AF). The navy-colored lines and text describe the second follow-up period.

#### Any Fever, Scheduled Visits

Before the first round of IRS (*i*.*e*., through November 2014), average baseline community fever prevalence was approximately 13.2% in the population, but it fell in the two post-IRS follow-up periods to 1% and 0.6% (Fig. 3). Pre-intervention community fever prevalence was highest in young children (18.4%), close to the average in older children (13%), and lowest in adults (8%). In the post IRS follow-up periods, community fever prevalence was highest in young children (1.6% and then 1.1%), lower in older children (1.2% and 0.7%) and lowest in adults (0.2% and 0.1%).

As an expected number of days in a year with fever, these proportions translate to 49 days *pppy* with community fever overall before IRS, and around 3 after: in young children, 67 before IRS and 4-6 after; in older children, 46 before and around 3 after; and in adults, 29 days with fever *pppy* before and around 0.5 after.

Similar patterns were also observed among patients who were parasite negative by LAMP or PCR (Fig. 3). In patients who were parasite negative by every test, community fever prevalence was 11.1% overall: 14.5% in young children; 11.5% in older children, and 7.9% in adults. After IRS, community fever prevalence among parasite negative individuals was 0.9 and 0.6% overall: 1.5% and 1% in young children; 0.7% and 1% in older children; and 0.2% and 0.1% in adults. In the before IRS *vs*. after IRS comparison, the MAFF was similar: 93-95% overall, 98-99% in adults, 90-95% in older children, and 91-94% in young children. Among patients who were also negative by LAMP and PCR (Fig. 3), the patterns were virtually identical: the MAFF was 92-95% overall, 98-99% in adults, 91-94% in older children, and 90-93% in young children.

#### Objective Fever, Scheduled Visits

Trends in community objective fever (*i*.*e*., during scheduled visits) broadly mirrored those of all fever, but prevalence of objective fever was much lower, and attributable fractions were also somewhat lower (Fig. S3). Overall baseline community objective fever prevalence was 2.4%, which declined to 0.5% and 0.4% in the two follow-up periods. Pre-IRS community prevalence of objective fever was highest in young children (4.4%), and lower in older children (2.3%). Objective fever was extraordinarily uncommon in adults; during the entire study (baseline and follow-up periods) only six objective fevers were observed during scheduled visits. In the follow-up periods objective fever prevalence was similar in young (0.7% and 0.5%) and older children (0.8% and 0.6%). The reduction in objective fever in the community was largest in young children (85% and 88%) and lower in older children (67% and 75%). Similar patterns were observed among patients who were parasite negative by LAMP or PCR in young children, but not in older children (Fig. S3). Among parasite negative patients, objective fever in the community was 0.8% overall: 1.5% in young children, 0.6% in older children. After IRS, prevalence of objective fever was 0.4-0.5% in young children, 0.5-0.6% in older children. Thus, the MAFF in young children was 64-72%; but in older children it was 0-6% (*i*.*e*., no reduction in fever was observed).

#### Fever and Patency, Scheduled Visits

The observed reduction in fever prevalence was accompanied by changes in infection prevalence by more sensitive measures and fever severity. There was a very large reduction in parasite prevalence by any metric (including submicroscopic infections), from 65% to 10-12% overall: from 60% to 4-5% in young children, 74% to 9-13% in older children, and from 52% to 10-11% in adults.(Nankabirwa et al., 2020) Figures 4 and 5 show the complex changes in parasitemia and fever before and after IRS. Fever prevalence among submicroscopic infections decreased from 14.1% to 1.4% in young children, 10.9% to 1.0% in older children, and 7.9% to 0.4% in adults, and among patients with low patent infection from 27.7% to 6.5% in young children, 15.6% to 3.7% in older children and 9.2% to 0% in adults.

**Fig. 4:**
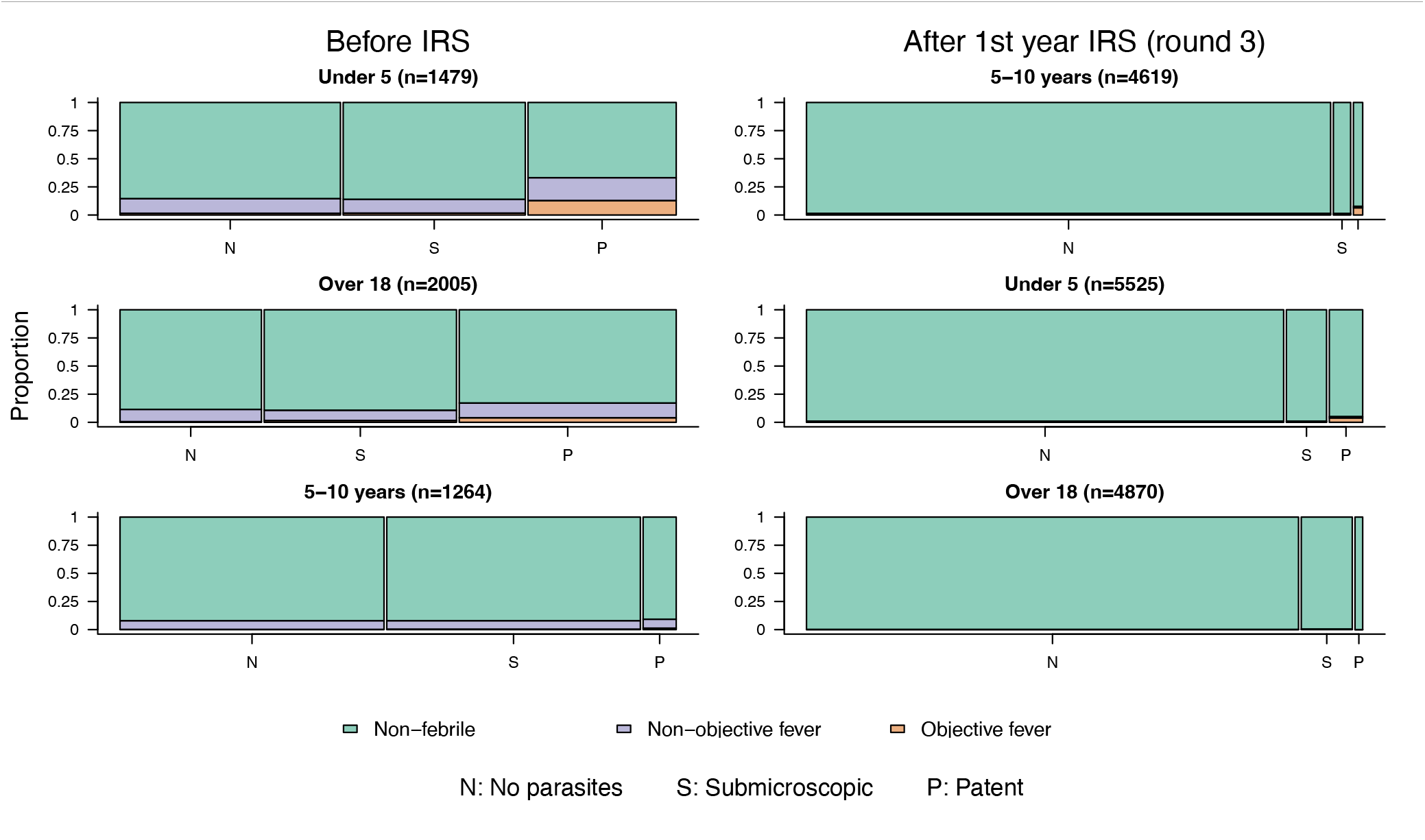
Fever and parasite status before and after IRS for scheduled visits. Widths of bars indicate the proportion of individuals in each parasite status category: no parasites (negative by light microscopy and LAMP/PCR), submicroscopic (negative by light microscopy and positive by LAMP/PCR) and patent (positive by light microscopy). The colors show the proportion of individuals with objective fever (temperature >38C), non-objective fever (subjective fever but temperature ≤38C), and without fever within each parasite status category. The columns compare the periods before IRS (left column) and after the end of the third round of IRS (right column). Results are plotted for young children (under 5), older children (5-10) and adults (over 18).

**Fig. 5:**
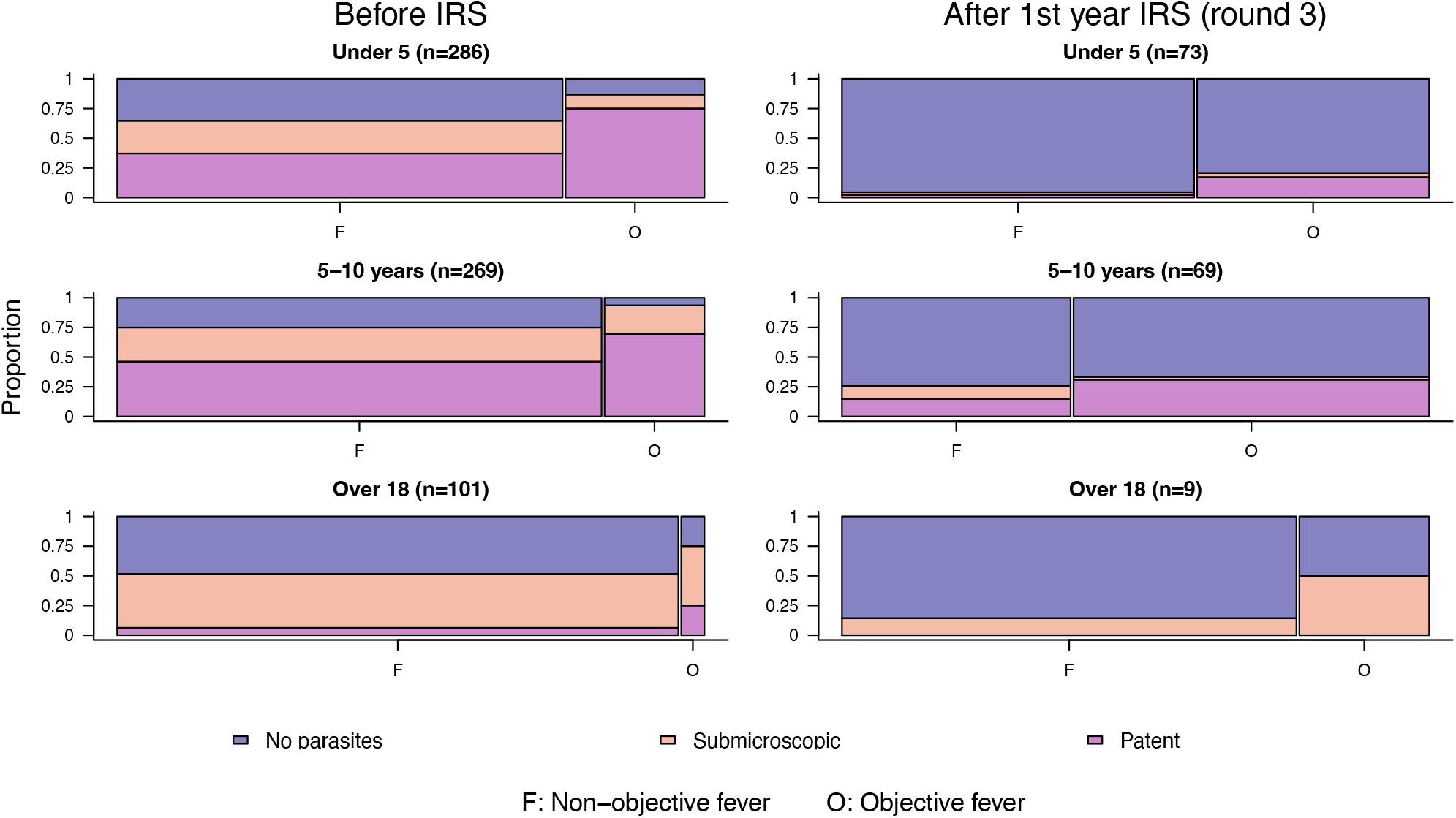
Parasites and fever before and after IRS for scheduled visits. The graphs compare parasite infection status and objective fever during scheduled visits for all febrile patients before and after the third round of IRS. Parasite status is shown stratified by the proportion of individuals with no detected parasites (*i*.*e*., negative by light microscopy and LAMP/PCR), submicroscopic (negative by light microscopy and positive by LAMP/PCR) and microscopically patent parasitemia (positive by light microscopy. The widths of bars indicate the proportion of febrile individuals with non-objective fever (subjective fever but temperature ≤38C) and objective fever (temperature >38C). Results are plotted for young children (under 5), older children (5-10) and adults (over 18).

**Fig. 6:**
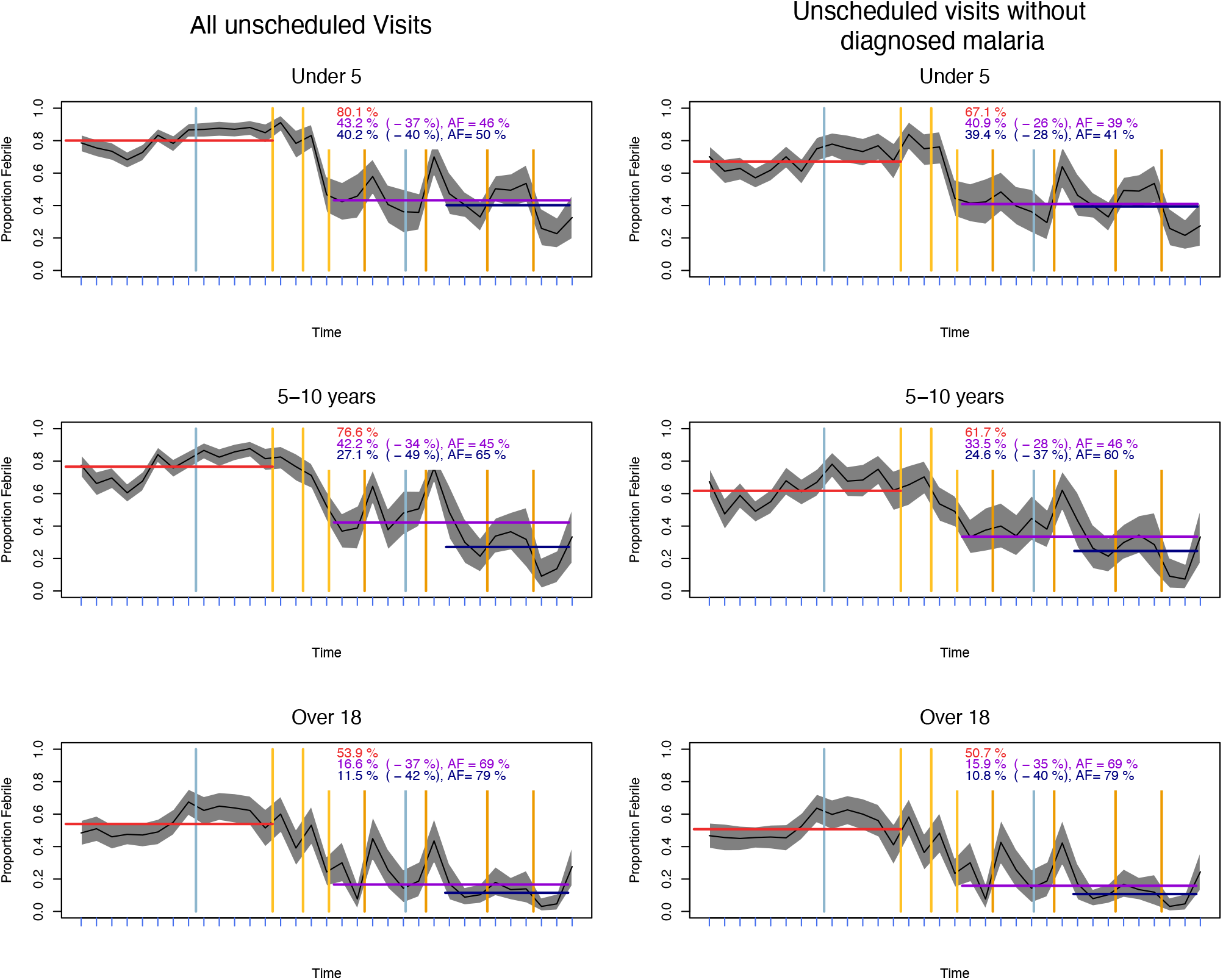
Clinical fever prevalence. The data were binned and plotted by quarter (blue tick marks) for all unscheduled visits (left column), and for a subset of unscheduled visits in which patients were diagnosed as malaria. The rows show young children (<5 years of age, top row), older children (5-10, middle row), and adults (>18, bottom row). The shaded grey regions show the binomial confidence intervals. Vertical yellow/orange lines indicate dates when spraying occurred, and blue lines are the dates of mass ITN distribution. The red horizontal line and red text describe community fever prevalence at the pre-IRS baseline. The purple horizontal lines and text describe community fever prevalence in the first follow-up period, the crude change in prevalence (in parenthesis), and the attributable fraction (AF). The navy-colored lines and text describe the second follow-up period.

Among febrile individuals, we observed changes in severity of fever (objective vs. non-objective, Fig. 5). The proportion of fevers in scheduled visits which were objective substantially increased in all age groups (from 24.3% to 40.5% in young children, 17.7% to 60.9% in older children, and 4.0% to 22.2% in adults). At the same time, there was a massive reduction in the proportion of febrile patients who had any parasitemia. In scheduled visits, parasite prevalence (including submicroscopic parasitemia) decreased from 60.0% to 16.2% in young children, 74.3% to 30.0% in older children and 52.1% to 17.5% in adults. Prevalence of patent parasitemia decreased from 24% to 2% in young children, from 36% to 6% in older children and from 6% to 1% in adults.

### Fever Prevalence during Unscheduled Visits

Clinical fever prevalence was defined herein as the proportion febrile during unscheduled visits for PRISM subjects. Here, we report the analysis for any clinical fever and objective clinical fever from the PRISM studies.

#### Any Fever, Unscheduled Visits

Clinical fever declined by approximately two-thirds overall (Fig. 7). In the PRISM studies before IRS, 73% of all unscheduled visits were accompanied by a fever. Clinical fever prevalence was highest in young children (80%), lower in older children (77%), and lowest in adults (54%). In the two follow-up periods after IRS, clinical fever prevalence fell to 33% and 26%: it was highest in young (43% and 40%), lower in older children (42% and 27%), and lowest in adults (17% and 12%). Among unscheduled visits not diagnosed with malaria, baseline fever prevalence was 60% overall: 67% in young children, 62% in older children, and 51% in adults. In the follow-up periods after IRS, fever prevalence among non-malaria clinical visits declined to 30% and 25% overall: 41% and 39% in young children; 34% and 25% in older children; and 16% and 11% in adults. Comparing fever in unscheduled visits before vs. after IRS, the fraction attributed to malaria was 54-64% overall, 46-50% in young children, 45-65% in older children, and 69-79% in adults. Among unscheduled visits not diagnosed with malaria, the MAFF was 51-59% overall, 39-41% in young children, 46-60% in older children, and 69-79% in adults.

**Fig. 7:**
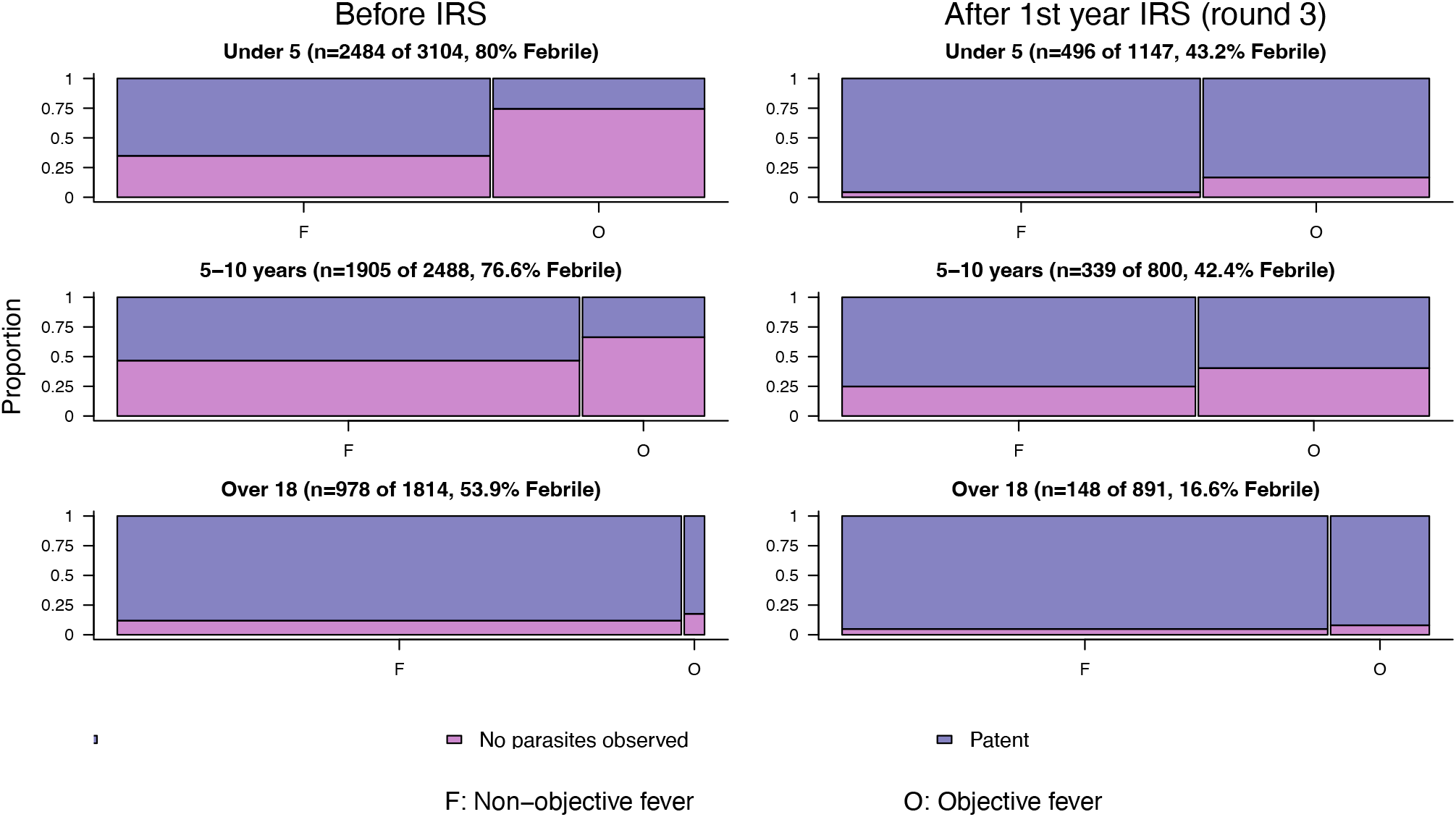
Parasites and fever before and after IRS for unscheduled visits. The graphs compare parasite infection status and objective fever among all febrile patients before and after the third round of IRS. Parasite status is shown stratified by the proportion of individuals with no detected parasites (*i*.*e*., negative by light microscopy), and microscopically patent parasitemia (positive by light microscopy). No LAMP/PCR data were collected during unscheduled visits. The widths of bars indicate the proportion of febrile individuals with non-objective fever (subjective fever but temperature <=38C) and objective fever (temperature > 38C). Results are plotted separately for young children (under 5), older children (5-10) and adults (over 18).

#### Objective Fever, Unscheduled Visits

We observed somewhat different patterns in objective fever prevalence during clinical visits. Before IRS 18% of all unscheduled visits were accompanied by an objective fever, which decreased only to 12% and 10% in the two follow-up periods. As with other indicators, objective fever prevalence in clinical visits was highest in young children (29%), lower in older children (16%), and lowest in adults (1.9%). In the two follow-up periods after IRS, prevalence of objective fever in clinical visits was highest in young children (16.7% and 16.2%), lowest in adults (2.8% and 2.9%), and decreased only in the second follow-up period in older children (16.7% and 12.6%). Notably, the MAFF for clinical objective fever prevalence followed a different pattern: it was 31-42% overall, 42-44% in young children, but much smaller in older children (−4-21%), and negative in adults (−50% and −53%) indicating an increase in objective fever prevalence among adults in clinical visits after IRS. However, except for the decrease in young children, these differences were not statistically significant (Table 1). Additionally, in contrast with all fever, there was no reduction of objective fever in unscheduled visits not diagnosed as malaria. In fact, there was a small but non-significant overall increase in the prevalence of objective fever in clinical visits not diagnosed as malaria from 7% before IRS to 10% and 9% after IRS: in young children prevalence increased from 12% to 15%, in older children from 9% to 11%, and in adults from 2% to 3%.

### Total Fever

To compute total fever, we would need to know the number of days with fever, per febrile clinical incident. It was not measured as part of the study, so we conducted a sensitivity analysis using values of *D* ranging from 1 up to 10 days (Fig. S6). Care seeking rates, which also affects total fever prevalence, also changed over the course of the study. Noting that overall disease incidence, as measured by care seeking rates (*i*.*e*., see Fig. 2), were changing over the whole course of the study, and that these patterns do not change in response to either IRS or to the changing protocol, we have estimated total fever in two ways.

First, to estimate the MAFF, we used the care seeking rates (*i*.*e*., *λ*) observed at the beginning of the study to estimate baseline clinical fever days: 5.8 visits *pppy* overall, 7.2 visits *pppy* for young children; 4 visits *pppy* for older children; and 5.6 visits *pppy* for adults. We report total fever assuming there were 2-4 days of fever, per incident (*i*.*e*., *D* = [2,4]), but also conducted a sensitivity analysis relaxing this assumption (Fig. S6). We estimated that there would be 57-65 days with fever *pppy* before IRS: 79-90 in young children; 54-60 in older children; and 35-41 in adults. Using the care seeking rates observed at the beginning of the study and fever prevalence from the first follow-up period, we estimate there would be 7-11 days *pppy* of total fever overall in the first follow-up period: 12-18 in young children, 8-11 in older children, and 3-4 in adults. In the second follow-up period, we estimate there would be 5-8 total fever days *pppy* overall: 10-16 in young children, 5-7 in older children, and 2-3 in adults. The MAFF was 83-87% overall in the first follow-up period: 80-85% in young children, 81-85% in older children, and 90-93% in adults. The MAFF in the second follow-up period was slightly higher, 87-90% overall: 83-87% in young children, 89-91% in older children, and 93-95% in adults.

Next, to estimate the total effect of the study, we used the care seeking rates observed at the end of the study to estimate fever in the follow-up periods: 1.4 visits *pppy* overall, 1.7 visits *pppy* in young children, 1 visit *pppy* in older children, and 1.6 visits *pppy* in adults. Using the care seeking rates observed at the end of the study and community and clinical fever prevalence from the 2nd follow-up period, we estimate that there would be 3-4 days *pppy* of total fever overall: 5-7 in young children, 3-4 in older children, and approximately 1 in adults. The overall difference in estimated total fever days at the start of the study compared to the end of the study was 94-95% overall: 92-93% in young children, 94-95% in older children, and 97-98% in adults.

### Fever in Outpatients, HMIS

We sought to understand whether the patterns we had observed in the PRISM studies were comparable to outpatient data for the same facility. We thus examined the UMSP data for members of the community who were not enrolled in the PRISM studies

The patterns for subjective fever in the community were like those observed in the longitudinal studies (Fig. 8). In the HMIS data, fever prevalence during care seeking declined by 64% overall: clinical fever prevalence in young children was 89% before IRS and 48-50% afterwards, a decline of approximately 64-65%; in older children, clinical fever prevalence declined by around 64% from 72% to 41%; in adults, clinical fever prevalence declined by 62-63% from 44% to 25-27%.

**Fig. 8:**
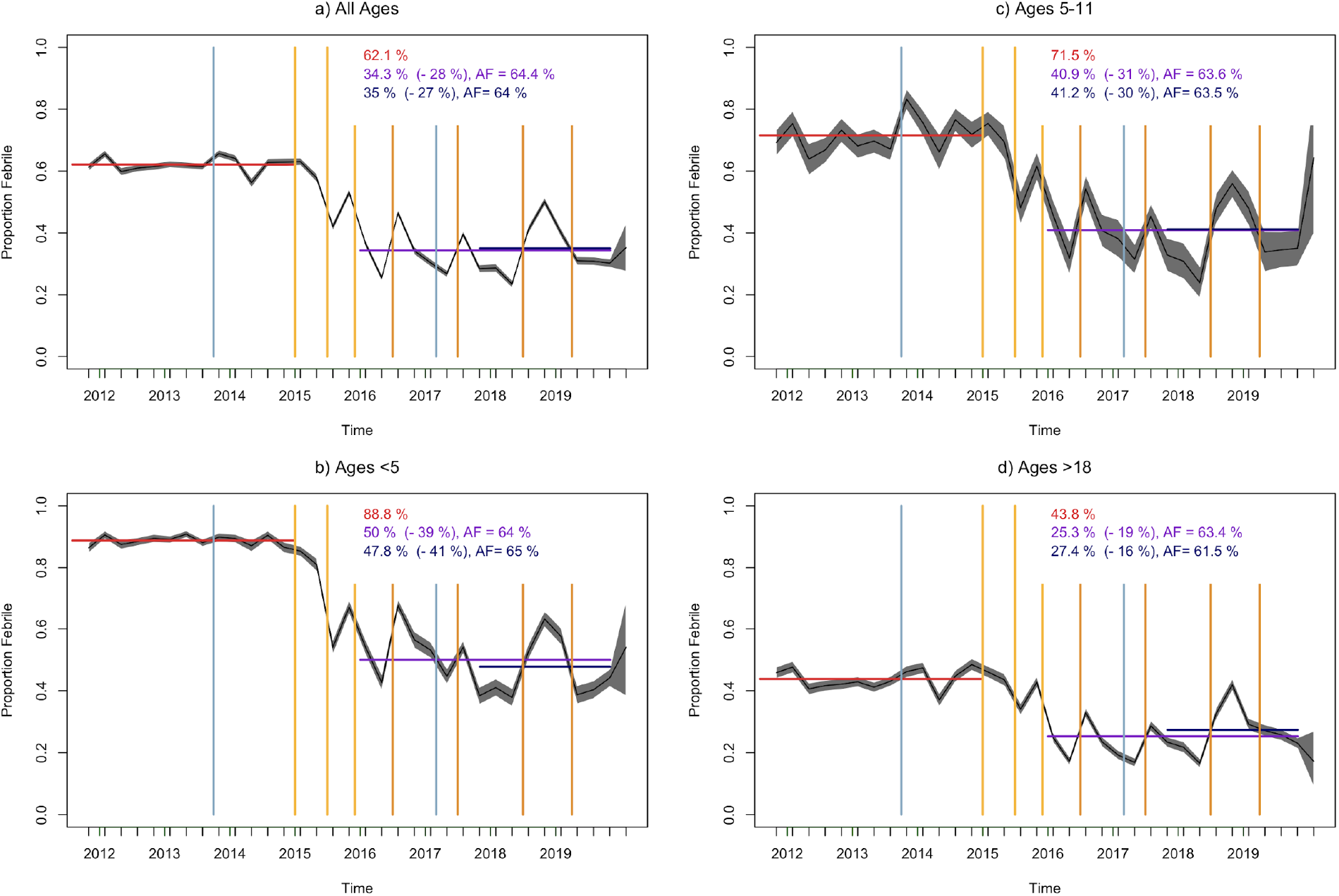
Clinical fever prevalence in UMSP data. The data and colors are identical to Fig. 5, but the study population is all patients reporting to the clinic in Nagongera, not including the PRISM study participants. As in Fig. 5, the data were binned and plotted by quarter and stratified by age. In this population, clinical fever prevalence differed by age but the fraction attributable to malaria was 62-65% across all ages.

## Discussion

These cohort studies in Nagongera, Tororo county, Uganda, were the first to measure the malaria attributable fever directly by making a comparison before and after malaria transmission was interrupted. Approximately 80-85% of total fever in children and 90-93% in adults was attributable to malaria. The study and our analysis suggest that subclinical disease (*i*.*e*., subjective fever that did not prompt an unscheduled visit to a clinic) was highly prevalent in this setting before the IRS spraying program, and that malaria was its dominant cause. Fever prevalence in the community declined by approximately 90-95% in children, and by 99% in adults. Notably, the reductions in fever in the community after IRS were observed across the population regardless of infection status. In subjects in the community with no detected parasites, fever was highly prevalent before IRS, but it was rare after IRS. Fever prevalence in the community was around 13% overall, but it was 11% in subjects who were parasite negative by light microscopy, LAMP, and PCR. After IRS, fever prevalence in the community dropped to below 1%. Malaria was also an important cause of fever with clinical disease, accounting for approximately two-thirds of fever in clinical visits in our cohort study: half of the fever in young children, 65% in children, and 80% in adults. Notably, over the same time period, fever prevalence during clinical visits declined by around 64% across all ages in patients reporting to the Nagongera clinic who were not enrolled in the study, as documented by the enhanced surveillance data (Sserwanga et al., 2011).

The observation that fever declined in the community, regardless of infection status, raises serious questions about the standard ways of defining malaria as a disease. In particular, the study draws attention to the enormous burden of malaria parasites as a cause of highly prevalent subclinical disease that is not associated with detectable parasitemia (Supplemental Figures). While there has been some debate about whether malaria is the cause of disease in outpatients with subjective fever who test positive for malaria parasites, this study has shown that there is a heavy burden of malaria attributable fevers in malaria-negative individuals. The gold standard for *aetiology* is, in the case of malaria, a comparison of disease with and without exposure to the bites of sporozoite positive mosquitoes, and by this standard, fever clearly declined, regardless of parasitemia.

The conclusion of this study is that malaria is a cause of disease in humans even when parasites are not detectable. As many studies have shown, the failure to detect parasites does not mean that parasites are not present. In this population, exposure to malaria (as measured by the aEIR) was very high, suggesting that parasites would be emerging from the liver every few days. At these high exposure rates before IRS, clinical fever prevalence in patients without diagnosed malaria followed the same trends as malaria incidence. After IRS, clinical fevers appeared to mirror human biting rates (Supplemental Figures). Since re-exposure to malaria was common in this setting before IRS, it is possible that subjective fever is an observable manifestation of an immune response that has actively suppressed infections emerging from the liver, an effect of persistent exposure to malaria even in the absence of patent blood stream infection. Lingering immunity and re-exposure could explain why fever mirrored entomological exposure after IRS. Our study thus suggests that the relationship between exposure to malaria and disease is complex, and that it may include indirect interactions among diseases through a range of mechanisms, such as chronic inflammation.

Notably, care seeking rates declined substantially over the course of the study, which could reflect a cumulative long-term health benefit that accrued over eight years of ready access to healthcare. Over the first year of the study, the frequency of care seeking (*i*.*e*., unscheduled visits) increased, a phenomenon that has been noticed in other settings when access to healthcare improves. After peaking, which occurred approximately one year into the study, the frequency of care seeking declined steadily so that care seeking rates were more than 75% lower at the end of the study compared with the beginning. The study has thus established reasonable expectations for fever prevalence in healthy, malaria-free populations in sub-Saharan African communities with good access to care. Our study showed that in the absence of malaria and with good access to health care, fever is rare: children under the age of five would average 5-7 days of fever per year, and adults would have less than 1 day of fever per year.

Our study also draws attention to important differences in fever and the MAFF that were found by age, by setting (*e*.*g*. clinic *vs*. community), and for objective *vs*. subjective fever. In addition to the other patterns, our study offers some relevant information about care seeking that could help in the design of future studies. First, we did not find that overall care seeking rates (*i*.*e*., unscheduled visits) changed substantially after the sharp reductions in exposure to malaria and malaria incidence, nor did we find that care seeking patterns were altered by a change in the frequency of scheduled visits. Second, and perhaps of greater importance, the fraction of clinical episodes for all causes that were accompanied by a fever declined by 50-60% after IRS, including fever in individuals not diagnosed with malaria. Our study thus suggests that constant exposure to malaria affects overall prevalence of fever, but that the indirect effects of malaria on total fever could be attributed to malaria through any simple diagnostic methods at the clinic.

Fever patterns and the estimated MAFF differed by age. Estimates of the MAFF were sensitive to both the denominator (*i*.*e*., fever attributed to all causes) and the numerator (*i*.*e*., fever attributed to malaria). Most studies that have estimated the MAFF have focused on objective fever in children. This study estimated that objective fever in the community declined by 88% in young children and 75% in older children, and objective fever during care seeking declined by 44% in young children. In other studies, reported estimates of the MAFF have usually fallen between 15-40%, though in some cases as high as 90% (Tabue et al., 2019). Notably, objective fever was much less common than subjective fever overall, and it was so rare in adults that it was not meaningful to study the changes. We found that the MAFF was much higher in adults. In adults, subjective fever in the community had fallen to around 0.1% by the end of the study (only nine adults reported subjective fever in the follow-up period), and objective fever was extremely rare. After IRS, fever prevalence was very low in adult populations, which explains why the MAFF was so high for adults: 99% of subjective fever in the community, and 79% of clinical subjective fevers were attributable to malaria. By way of contrast, fever prevalence remained comparatively high in children after IRS, which explains partly why the MAFF was lower. Community fever prevalence declined by 90-95% in children of all ages, but there were larger differences in the MAFF for clinical fever prevalence: 50% in young children *vs*. 65% in older children. For objective fevers, age-differences for the fraction of fever attributable to malaria in the community were broadly similar, though perhaps lower in older children than young children or adults.

Fever and malaria are important causes of disease in sub-Saharan Africa, but studies of fever etiology have rarely found that malaria is one of many causes of clinical fevers (D’Acremont et al., 2014). Much of the previous work on malaria attributable fever fraction has been motivated by a need to define thresholds for clinical diagnosis that are optimally sensitive and specific (Schellenberg et al., 1994). Indeed, many authors have argued that relatively low malaria attributable fever fractions suggest overdiagnosis and overtreatment of malaria could be problematic in clinical fever management. This study casts substantial doubt on such an argument, though the implications of a very high MAFF on clinical fever management and parasitemia-based case definitions are not clear (Smith et al., 1995). While we have found a very large malaria attributable burden of fever, much of this burden occurred in individuals without detectable parasitemia. The most relevant comparison from before and after IRS was that the prevalence of fever during care seeking declined substantially in both malaria positive and malaria negative individuals, and that this was observed in the research study as well as the surveillance data from the community. Malaria fever was a complicating factor in the diagnosis and treatment of more than half of all clinical visits.

One important caveat is that this study was not designed to measure attributable fever as an outcome of the study, and the onset of IRS unfortunately coincided with a change in the study protocol, when scheduled visits for children changed from once every three months to once a month. It is possible that some of the effects we have attributed to malaria are, in fact, attributable to a change from quarterly to monthly visits to a clinic, though the changing patterns of scheduled visits did not exactly follow the protocol (Fig. 3). We note that the protocol changed at different points in time for adults and children, and that the changes in fever in adults followed IRS, not the change in study protocol. Acknowledging that uncertainty about whether IRS or the study protocol caused the changes cannot be resolved by this study, we have thus attributed the changes in fever to the absence of malaria, but with some caution. A new study would be required to address this major caveat.

A second notable caveat is that malaria was not completely absent after IRS, so it could not be completely ruled out as a cause of fever. After IRS, exposure was rare and during the last two years of the study, exposure had dropped to less than one infection, per person, per year. We note that baseline exposure to malaria in this study population was high; individuals were expected to be infected, on average, more than once per week. Because of ongoing exposure to malaria after IRS, some fever in the follow-up could be causally related to malaria. Indeed, there appears to be an association between the HBR and fever in unscheduled visits after IRS (Figure S5). If so, we may have under-estimated the MAFF.

A third caveat of this study is that it is not possible to know what effects IRS may have had on the transmission of non-malarial pathogens causing fever. Given that IRS involves only indoor spraying, any additional effects would likely be on other vector-borne pathogens transmitted by indoor-biting arthropods. Previously, unbiased sequencing of serum from a convenience sample of febrile children in Tororo District Hospital found a range of pathogens present, but none of those transmitted by arthropods were present in nearly the same magnitude of patients as *P. falciparum* (Ramesh et al., 2019). Indeed, a substantial portion of malaria-positive serum was coinfected with other pathogens, and the study identified at least one possible important interaction between malaria and parvovirus B19 as a cause of fever. This supports our assumption that any reductions in transmission of other pathogens because of IRS had a small effect on fever rates and suggests that interactions between malaria and other pathogens could be an important mechanism by which chronic exposure to malaria causes fever.

This study has provided an opportunity for the first time to directly measure the malaria attributable fraction of fever in a highly endemic area, and in a subsequent manuscript, we plan to draw comparisons by using these same data to estimate the MAFF using other methods. The trends in this study paint a substantially different picture of the burden of malaria than has previously been described. After IRS, subjective and objective fever prevalence were substantially reduced, but there were proportionally more objective fevers; most malarial disease was not especially severe overall in this study, but mild disease is very common. While some previous studies have estimated malaria attributable fever among adults (Afrane et al., 2014), we have found a much higher attributable burden of disease, which was mostly subjective fever caused by malaria. More broadly, these findings point to the importance of subjective fever as an outcome to consider in the full burden of malaria in all age groups. Indeed, the main implication of this study is that there appears to be a sizable burden of malaria-attributable fever not associated with measurable parasitemia. It is possible the increased burden of fever associated with malaria is also associated with severe disease and poor outcomes for other conditions. Other evidence suggests malaria is also a major cause of death, and there is also a heavy burden associated with severe malaria. If patterns hold throughout highly endemic regions, removing malaria would probably have large direct and indirect benefits: direct effects would include removing malaria as a cause of malaria death and severe malaria; and indirect benefits would include removing the mediating effect of malaria as a cause of increased death and severe disease from other causes. In areas where malaria is highly prevalent, removing malaria is arguably the single most important approach to improving overall health and will likely have benefits beyond the expectations of most experts.

## Data Availability

All data from these studies are publicly available at ClinEpiDB.

https://clinepidb.org/ce/app/workspace/analyses/DS_51b40fe2e2/new

https://clinepidb.org/ce/app/workspace/analyses/DS_0ad509829e/new/details.

## Supplemental Figures

**Fig. S1:**
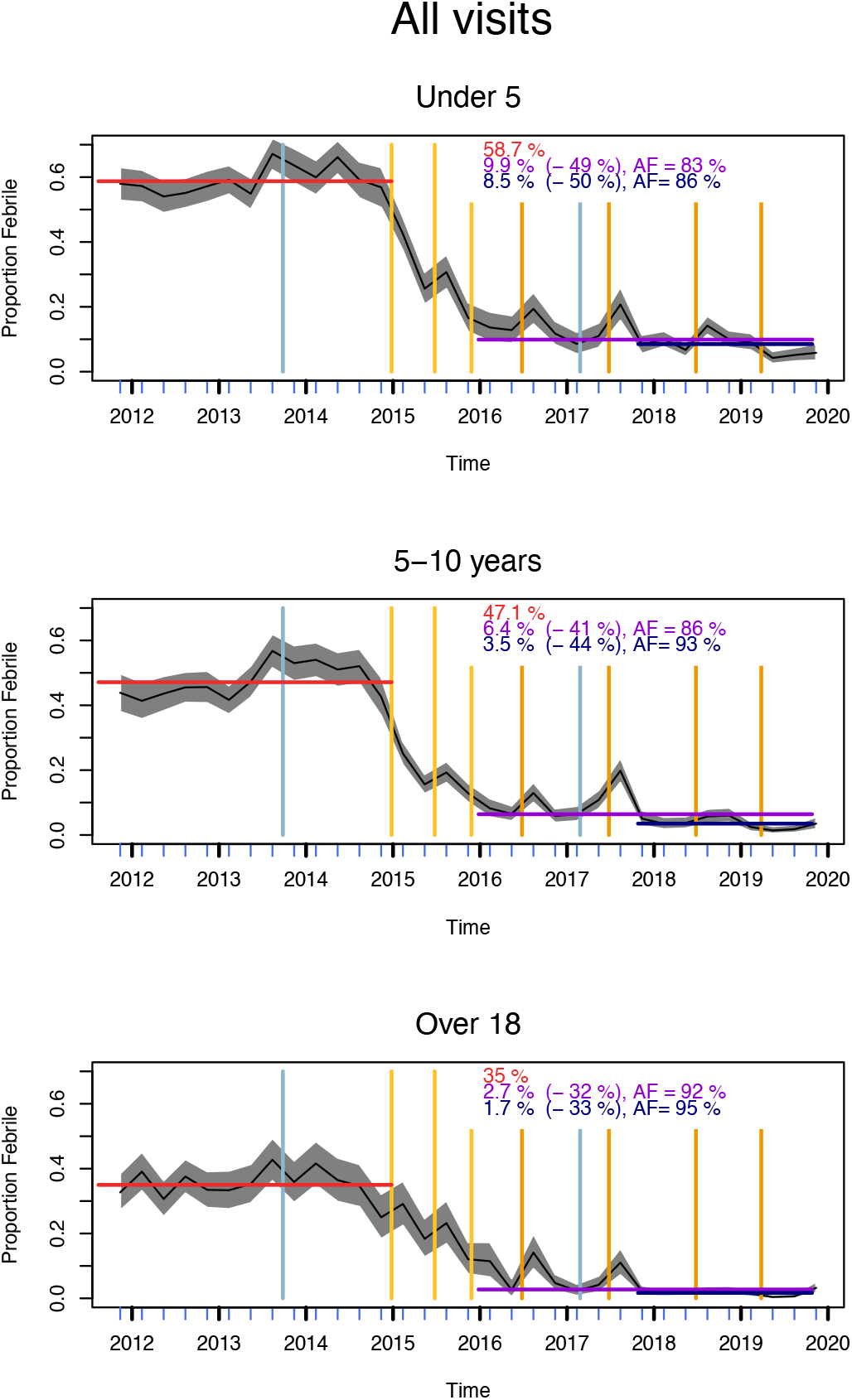
Total fever prevalence over all observations during the study. Results are plotted by quarter (blue tick marks) for a) all study subjects; b) young children; c) older children; and d) adults. The shaded grey regions show the binomial confidence intervals. Vertical yellow/orange lines indicate dates when spraying occurred, and blue lines are the dates of mass ITN distribution. The red horizontal line and red text describe fever prevalence among all observations at the pre-IRS baseline. The purple horizontal lines and text describe prevalence in the first follow-up period, the crude change in prevalence (in parenthesis), and the attributable fraction (AF). The navy-colored lines and text describe the second follow-up period.

**Fig. S2:**
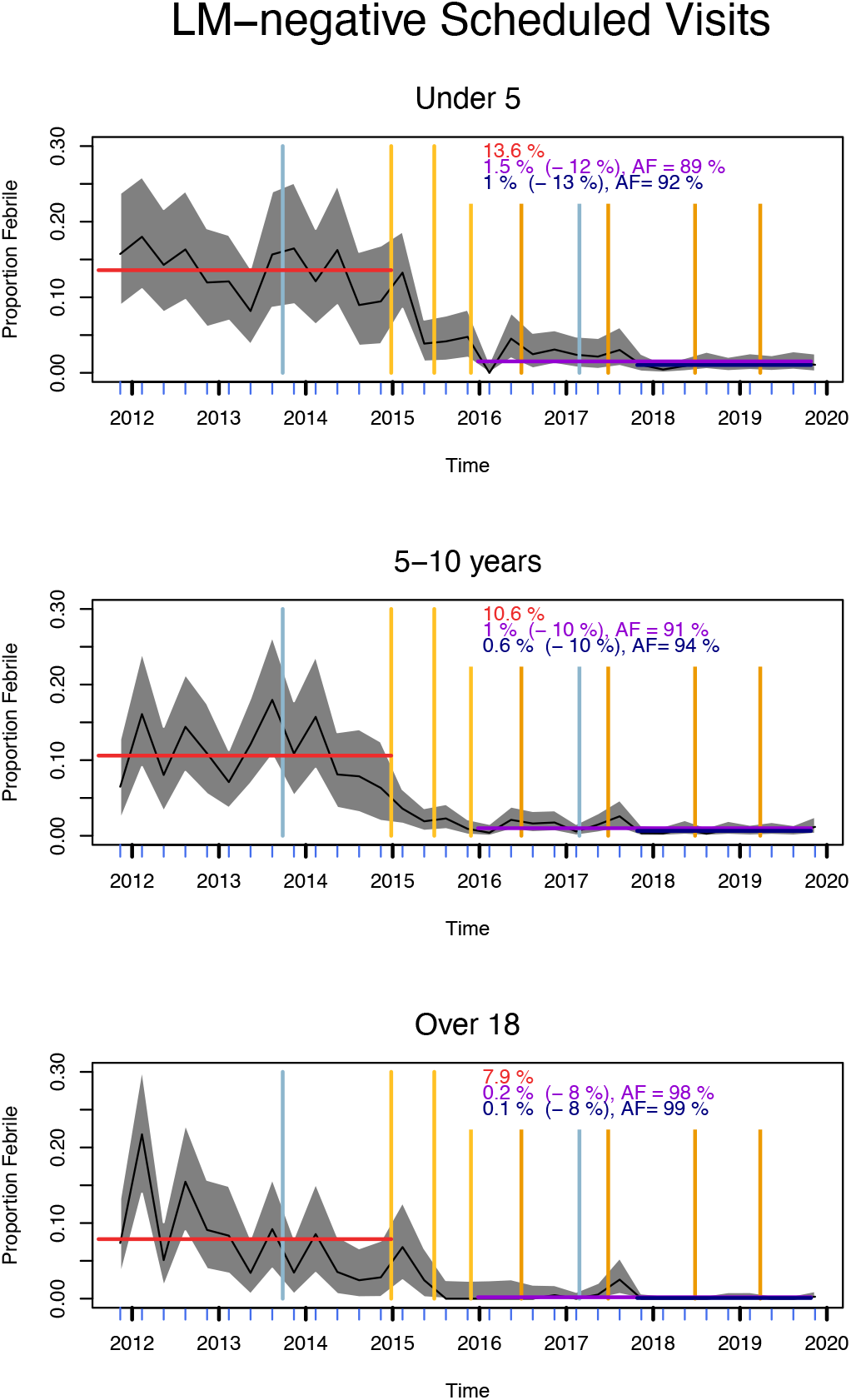
Community fever prevalence among non-patent (negative by light microscopy) individuals as observed during the study. Note that parasite density was measured during all scheduled visits. Results are plotted by quarter (blue tick marks) for a) all study subjects; b) young children; c) older children; and d) adults. The shaded grey regions show the binomial confidence intervals. Vertical yellow/orange lines indicate dates when spraying occurred, and blue lines are the dates of mass ITN distribution. The red horizontal line and red text describe community fever prevalence at the pre-IRS baseline. The purple horizontal lines and text describe community fever prevalence in the first follow-up period, the crude change in prevalence (in parenthesis), and the attributable fraction (AF). The navy-colored lines and text describe the second follow-up period.

**Fig. S3:**
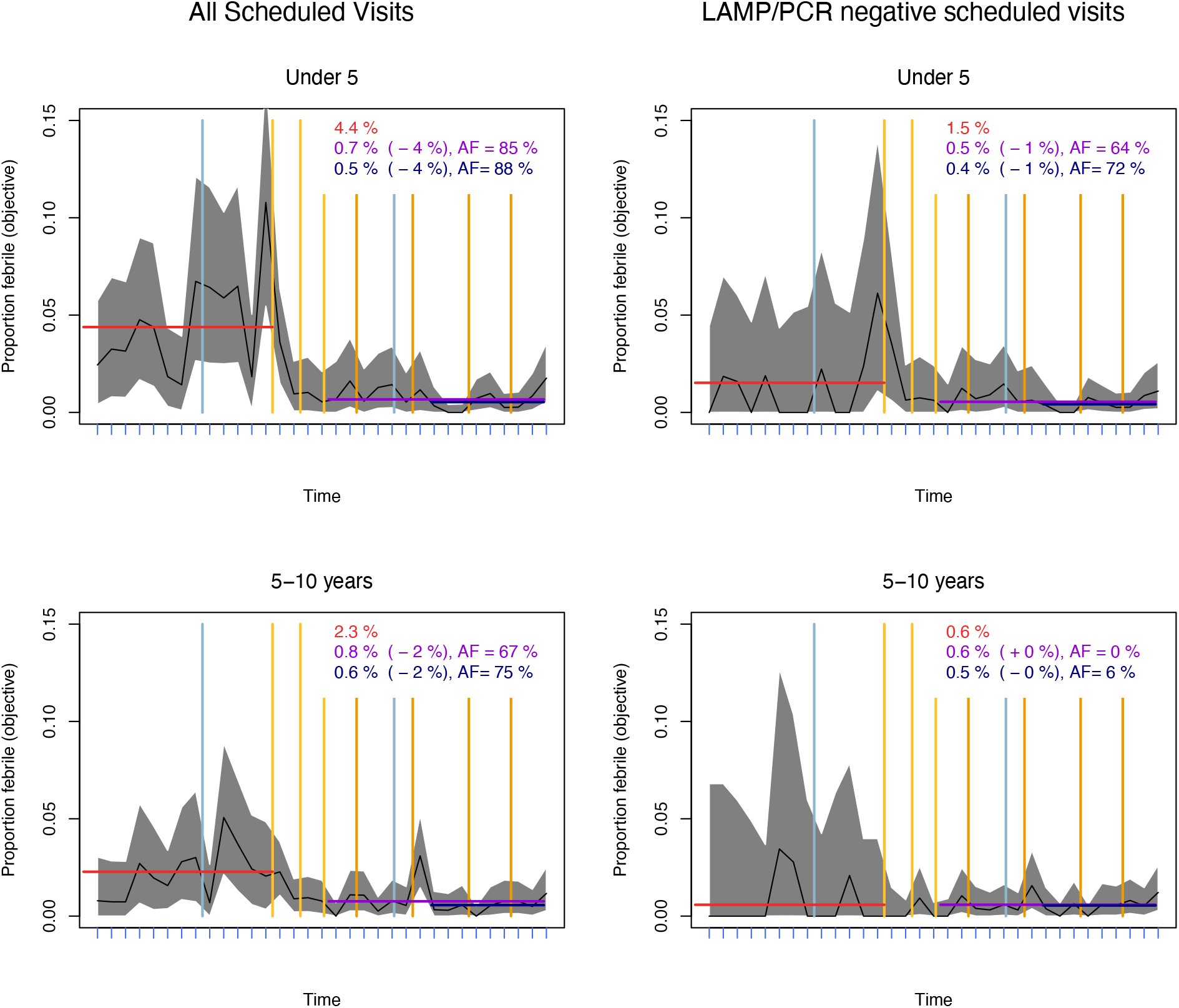
Community objective fever prevalence. plotted by quarter (blue tick marks) for all scheduled visits (left) and only scheduled visits malaria-negative by LAMP (PRISM 1) or PCR (PRISM 2), in young children (top row), older children (bottom row). Only six objective fevers in adults during scheduled visits were observed during the study, so analogous calculations are not meaningful in this group. Note that LAMP or PCR was conducted for 98% of scheduled visits. The shaded grey regions show the binomial confidence intervals. Vertical yellow/orange lines indicate dates when spraying occurred, and blue lines are the dates of mass ITN distribution. The red horizontal line and red text describe community fever prevalence at the pre-IRS baseline. The purple horizontal lines and text describe community prevalence in the first follow-up period, the crude change in prevalence (in parenthesis), and the attributable fraction (AF). The navy-colored lines and text describe the second follow-up period.

**Fig. S4:**
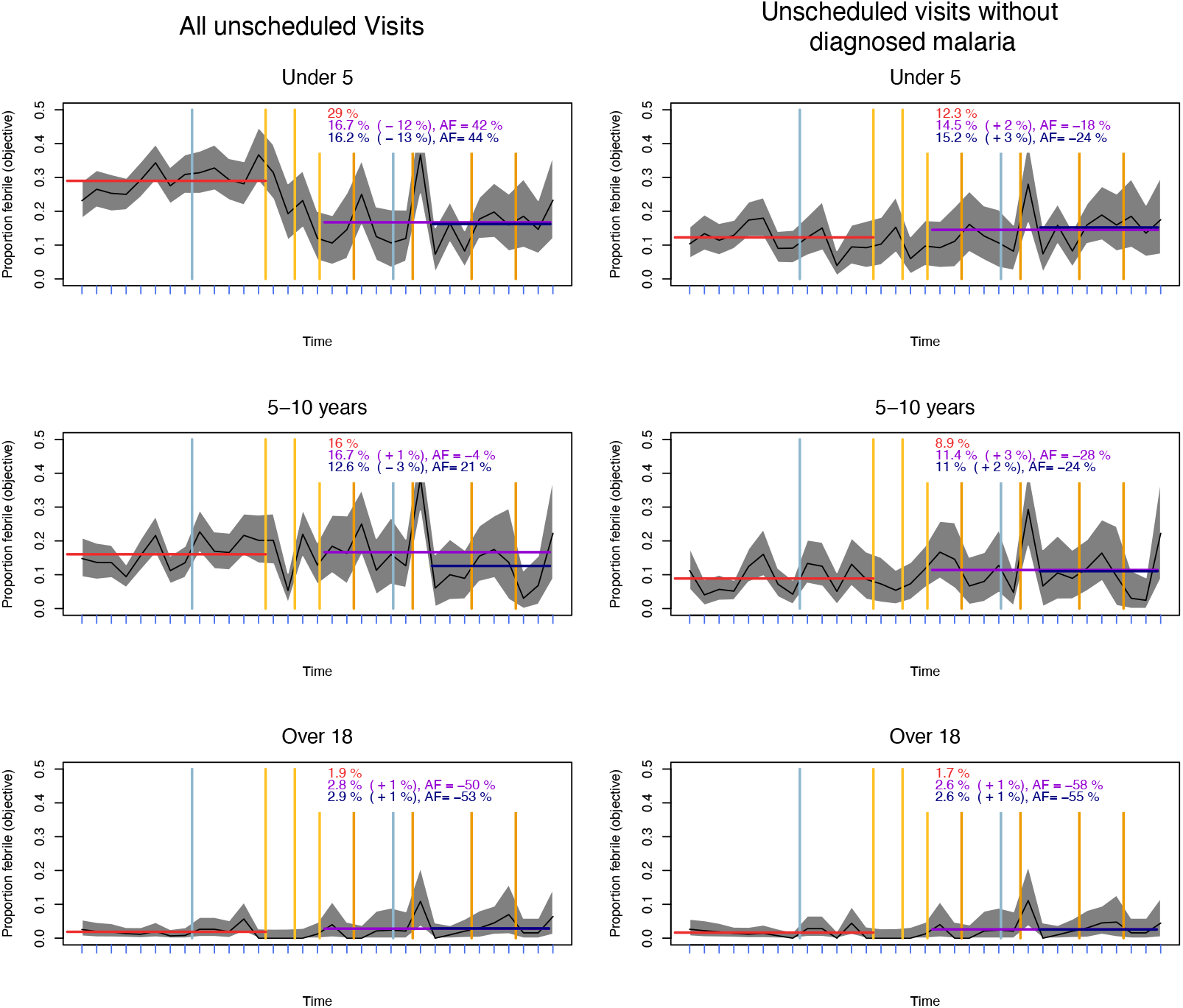
Clinical objective fever prevalence. plotted by quarter (blue tick marks) for all unscheduled visits (left) and only unscheduled visits not diagnosed as malaria, in young children (top row), older children (middle row), and adults (bottom row). Note that all febrile patients had a blood smear taken, and those positive by light microscopy were diagnosed with malaria, but parasitemia measurements were not made for many non-febrile patients. The shaded grey regions show the binomial confidence intervals. Vertical yellow/orange lines indicate dates when spraying occurred, and blue lines are the dates of mass ITN distribution. The red horizontal line and red text describe clinical fever prevalence at the pre-IRS baseline. The purple horizontal lines and text describe clinical prevalence in the first follow-up period, the crude change in prevalence (in parenthesis), and the attributable fraction (AF). The navy-colored lines and text describe the second follow-up period.

**Fig. S5:**
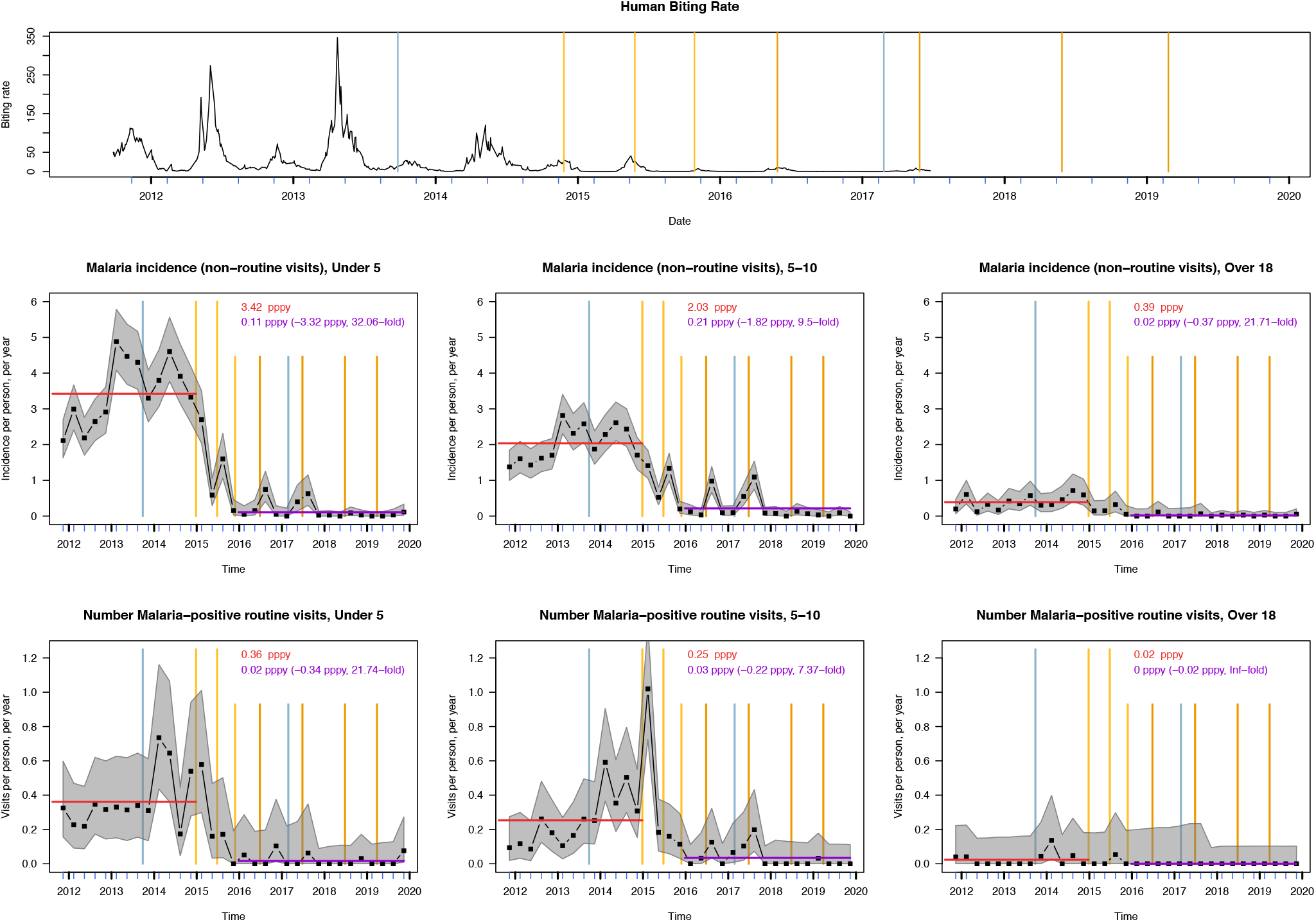
Changes in the human biting rate (top row), malaria incidence in young children (middle left), older children (middle center), and adults (middle right) and number malaria-positive scheduled visits in young children (bottom left), older children (bottom center), and adults (bottom right).

**Fig. S6:**
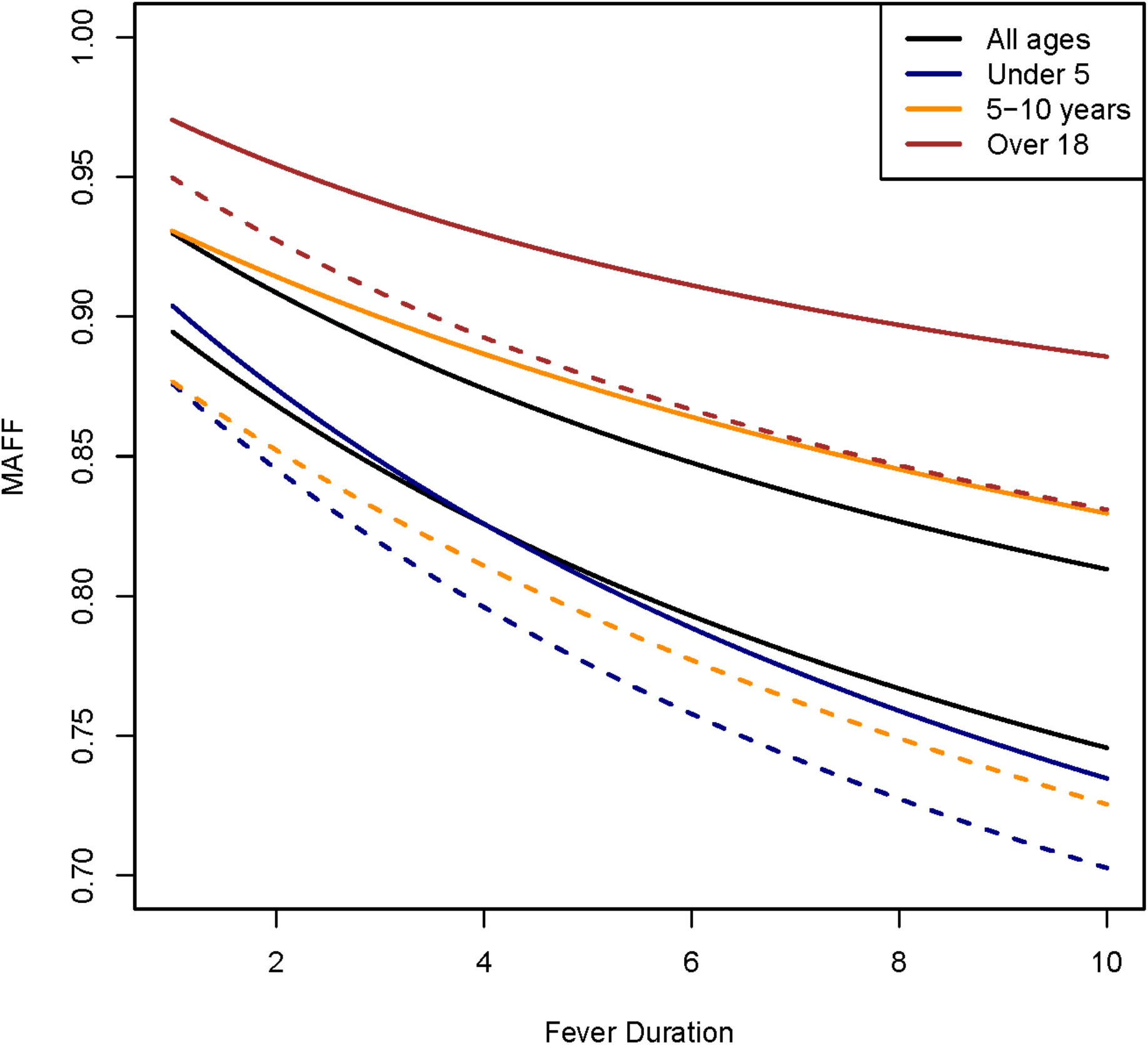
Sensitivity analysis of attributable fraction of total fever days using different fever durations. Estimated MAFF plotted against fever duration in days for all ages (black), young children (blue), older children (orange), and adults (red). MAFF for follow-up period 1 (from the end of round three of IRS to the end of the study) are shown in dashed lines, and for follow-up period 2 (the final two years of the study) in solid lines. All estimates assumed constant care-seeking at the level observed at the beginning of the study in each age group.

